# Surgical outcomes in complicated appendicitis: does timing or surgeon seniority matter? A propensity score-matched analysis from the RIFT Turkey cohort

**DOI:** 10.64898/2026.05.19.26353556

**Authors:** Ali Yalcinkaya, Semra Demirli Atici, Cihan Ozen, Deniz Karasoy, Erdinc Kamer, Ahmet Yalcinkaya, Sezai Leventoglu, RIFT Turkey Study Collaborators

**Affiliations:** Department of General Surgery, Faculty of Medicine, Gazi University, Ankara, Turkey; Department of Clinical Medicine, Faculty of Medicine, Aalborg University, Aalborg, Denmark; Department of General Surgery, Acıbadem Kent Hospital, Izmir, Turkey; Department of Gastrointestinal Surgery, Aalborg University Hospital, Aalborg, Denmark; Department of Medicine, Holbaek Hospital, Holbaek, Denmark; Department of General Surgery, Izmir Faculty of Medicine, University of Health Sciences, Izmir, Turkey; Department of Medical Biochemistry, Faculty of Medicine, Hacettepe University, Ankara, Turkey; Department of Medical Biochemistry and Microbiology, Science for Life Laboratory, Uppsala University, Uppsala, Sweden

**Keywords:** complicated appendicitis, surgical timing, night surgery, weekend effect, surgeon seniority, propensity score matching, RIFT Turkey

## Abstract

**Background:** Complicated acute appendicitis carries a higher risk of postoperative morbidity relative to uncomplicated cases. It remains unclear whether surgical timing (night vs. day; weekend vs. weekday) or surgeon seniority influence short-term outcomes in this high-risk population.

**Methods:** This was a retrospective analysis of the RIFT Turkey dataset restricted to histologically confirmed cases of complicated appendicitis who had undergone laparoscopic appendectomy. Primary exposures were surgical timing (day [n=92] vs. night [n=123]; weekday [n=172] vs. weekend [n=43]) and surgeon seniority (trainee [n=89] vs. consultant [n=126]). The primary outcome was unplanned readmission and/or reintervention within 60 days. Secondary outcomes were conversion to open surgery and length of stay (LOS) >3 days. Propensity score matching (PSM) using RIPASA score (caliper 0.05, SMD <0.1) was performed as a pre-specified sensitivity analysis for each comparison.

**Results:** Night-time surgery was associated with higher frequencies of unplanned readmission / reintervention (12.2% vs. 6.5%; OR 1.99 [95% CI 0.74-5.35], p=0.166) and surgical conversion (9.8% vs. 3.3%; OR 3.21 [0.88-11.72], p=0.064) compared with daytime surgery, neither reaching significance. Trainee surgeons had significantly higher readmission/reintervention rates than consultants (15.7% vs. 5.6%; OR 0.32 [0.12-0.82], p=0.013). PSM-adjusted results also showed similar relationships: night vs. day (readmission OR 2.45 [0.85-7.03], p=0.09; conversion OR 2.84 [0.73-11.1], p=0.13), weekend vs. weekday (readmission OR 1.53 [0.24-9.72], p=0.65), and trainee vs. consultant (readmission OR 0.25 [0.08-0.79], p=0.013).

**Conclusion:** Surgical timing was not significantly associated with short-term outcomes in complicated appendicitis, though night-time surgery showed a consistent trend towards higher complication rates. Surgeon seniority was the only factor independently and significantly associated with unplanned readmission and reintervention in both primary and PSM analyses, indicating the need for senior supervision during out-of-hours procedures.

## Introduction

Acute appendicitis is one of the most common surgical emergencies worldwide [1,2]. Although most cases are treated easily through well-established approaches, cases with complicated appendicitis are at risk for major complications that can increase risks for morbidity and mortality, including perforation, gangrenous change, periappendiceal abscess, or diffuse peritonitis. Complicated appendicitis also causes greater surgical difficulty and may lead to higher complication rates and increased resource utilization compared to uncomplicated disease [3,4].

Emergency surgical departments operate continuously, and appendectomies are frequently performed outside standard working hours. The concept of the ‘weekend effect’, whereby patients admitted at weekends experience worse outcomes than those on weekdays, has been described across multiple acute conditions [5]. Similarly, concerns exist about the quality of night-time surgical care, driven by potential differences in staffing, fatigue, and reduced specialist support [6]. For acute appendicitis, however, the existing evidence is inconsistent. A recent systematic review and meta-analysis by Hu et al. found that out-of-hours appendectomy was associated with higher conversion rates but no significant increase in postoperative complications or mortality [7]. Other studies synthesizing evidence from thousands of patients also suggest that the time of day during index surgery has no impact on surgical conversion, length of stay (LoS), readmission, or complications [8][9].

In parallel, surgeon seniority is also understood as being a determinant of surgical outcomes. Appendectomy is one of the first independently performed emergency procedures for trainees in general surgery departments and the abilities in perform appendectomies has been used as a barometer of training competency [10]. A systematic review and meta-analysis by Anyomih et al. found no significant difference in morbidity, conversion, or blood loss between trainees and trained surgeons performing appendectomy, though trainees had longer operating times [11]. However, this reassuring finding was derived from surgeries performed in mixed appendicitis populations. Complicated appendicitis, characterized by greater intraoperative complexity, may represent a setting where the margin for error is narrower, and where trainee-led surgery without adequate supervision could carry higher risks [12]. Furthermore, surgical timing and surgeon seniority may interact, particularly since night-time cases are often delegated to less-experienced surgeons operating with reduced senior oversight, potentially compounding the independent effects of both exposures. Clarifying the underlying relationships necessitates focused analyses based on prospectively collected data sets in which both variables are assessed concurrently and reliably in multiple centers.

The Right Iliac Fossa Treatment (RIFT) Turkey study is a nationwide multicentre prospective cohort that has previously characterized the epidemiology and diagnostic performance of appendicitis risk prediction models across Turkey [13,14]. It is a unique resource with highly accurate and data collection. The present study leverages this dataset, restricting analysis to patients with histologically confirmed complicated appendicitis undergoing planned laparoscopic appendectomy. Our purpose was to evaluate the independent associations of surgical timing and surgeon seniority with short-term postoperative outcomes, also utilizing propensity score matching as a pre-specified sensitivity analysis.

## Methods

### Study design and data source

This is a retrospective secondary analysis of the RIFT Turkey dataset, a nationwide multicentre prospective observational cohort study conducted across 84 centres in Turkey between September and December 2020. The design, ethical approval, patient eligibility criteria, data collection methodology, and primary outcomes of the parent study have been reported in detail elsewhere [13]. Briefly, all consecutive adults presenting with acute right iliac fossa pain or suspected appendicitis were prospectively enrolled. Data were collected using standardized case report forms and transferred to a central online database on REDCap, followed by rigorous quality control of data and data sources [15,16]. Ethical approval was obtained from the Clinical Research Ethics Committee of Gazi University Faculty of Medicine (7 September 2020), and the had been study registered on ClinicalTrials.gov (NCT04614649). All patients provided written informed consent. The study was conducted in accordance with the Helsinki Declaration.

### Study population

From the full RIFT Turkey cohort (n=3,358), we included patients if they met the following two criteria: (1) histologically confirmed complicated appendicitis (gangrenous or perforated appendicitis on pathological examination) and (2) initial operative approach planned as laparoscopic (including cases that subsequently converted to open surgery). Patients undergoing primary open appendectomy were excluded to ensure comparability of the operative context. Patients with missing operative time data were also excluded.

### Exposures

Three pre-specified exposures were evaluated. The first two concerned surgical timing: (i) time of day, dichotomized as daytime (08:00-16:59) versus night-time (all other hours); and (ii) day of the week, dichotomized as weekday (Monday-Friday) versus weekend (Saturday-Sunday). The third exposure was surgeon seniority, dichotomized as trainee (residents, senior residents, and registrars) versus consultant/attending surgeon. All data were prospectively recorded at the time of data collection.

### Outcomes

The primary outcome was presence of any unplanned readmission and/or reintervention within 60 days of the index procedure, including readmission to hospital, radiological drainage, and/or reoperation. Complications requiring readmission or reintervention were classified according to the Clavien-Dindo Classification [17]. Secondary outcomes were intraoperative conversion from laparoscopic to open surgery and prolonged length of stay, defined as LOS >3 days.

### Statistical analysis

Continuous variables are presented as median [interquartile range (IQR)] and categorical variables as frequencies and percentages. Baseline comparisons were performed using the Mann-Whitney U test for continuous variables and Pearson’s chi-square test or Fisher’s exact test for categorical variables, as appropriate. Unmatched outcome comparisons were performed using Pearson’s chi-square test or Fisher’s exact test, as appropriate, and effect estimates are presented as unadjusted odds ratios (ORs) with 95% confidence intervals (CIs). Due to the limited number of outcome events, multivariable analyses were not performed.

Propensity score matching (PSM) was performed as a pre-specified sensitivity analysis for each comparison. Propensity scores were estimated using logistic regression, with the RIPASA score included as a parsimonious summary measure of baseline disease severity, as this score has previously shown the best performance in this population [13]. One-to-one nearest-neighbour matching without replacement was applied using a caliper of 0.05 on the logit scale. Balance was assessed using the standardised mean difference (SMD), with SMD <0.1 considered indicative of adequate balance. In the matched cohorts, categorical outcomes were compared using Pearson’s chi-square test or Fisher’s exact test, as appropriate, and effect estimates are presented as ORs with 95% CIs. All analyses were performed using SPSS version 25 (IBM Corp., Armonk, NY, USA). A two-tailed p<0.05 was considered statistically significant.

## Results

### Cohort derivation and baseline characteristics

Of 3,358 patients enrolled in the RIFT Turkey parent study, 449 had histologically confirmed complicated appendicitis. After excluding 234 patients who underwent primary open appendectomy and 8 patients with missing operative time data, 215 patients formed the final analytic cohort. Of these, 123 (57.2%) underwent night-time surgery and 43 (20.0%) had weekend surgery. Trainee surgeons performed 89 (41.4%) of procedures. Baseline characteristics are shown in Table 1.

**Table 1.**
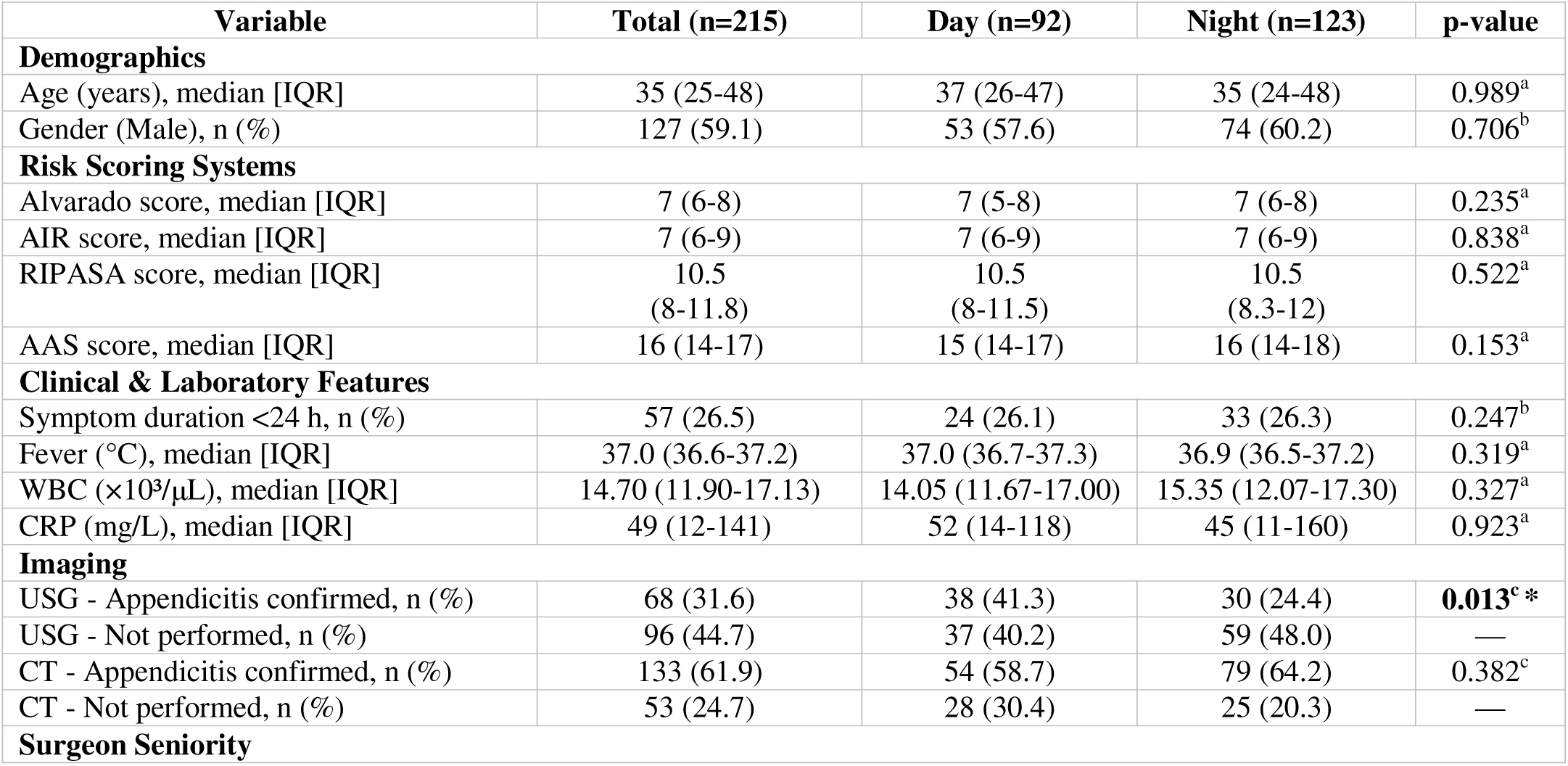
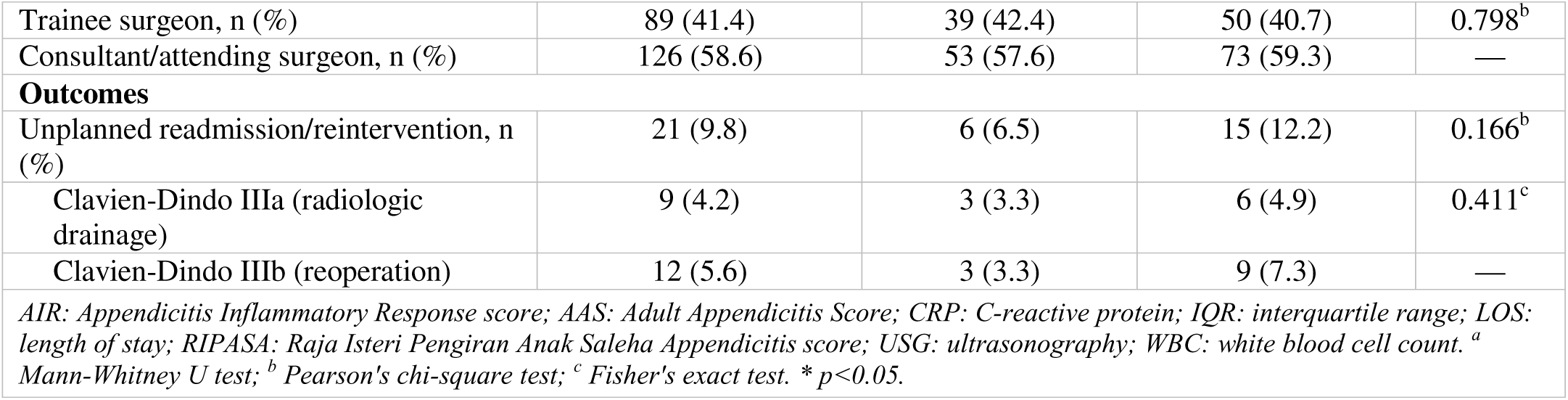
Baseline characteristics of the study cohort stratified by time of surgery.

| Variable | Total (n=215) | Day (n=92) | Night (n=123) | p-value |
| --- | --- | --- | --- | --- |
| **Demographics** |  |  |  |  |
| Age (years), median [IQR] | 35 (25-48) | 37 (26-47) | 35 (24-48) | 0.989<sup>a</sup> |
| Gender (Male), n (%) | 127 (59.1) | 53 (57.6) | 74 (60.2) | 0.706<sup>b</sup> |
| **Risk Scoring Systems** |  |  |  |  |
| Alvarado score, median [IQR] | 7 (6-8) | 7 (5-8) | 7 (6-8) | 0.235<sup>a</sup> |
| AIR score, median [IQR] | 7 (6-9) | 7 (6-9) | 7 (6-9) | 0.838<sup>a</sup> |
| RIPASA score, median [IQR] | 10.5<br>(8-11.8) | 10.5<br>(8-11.5) | 10.5<br>(8.3-12) | 0.522<sup>a</sup> |
| AAS score, median [IQR] | 16 (14-17) | 15 (14-17) | 16 (14-18) | 0.153<sup>a</sup> |
| **Clinical & Laboratory Features** |  |  |  |  |
| Symptom duration <24 h, n (%) | 57 (26.5) | 24 (26.1) | 33 (26.3) | 0.247<sup>b</sup> |
| Fever (°C), median [IQR] | 37.0 (36.6-37.2) | 37.0 (36.7-37.3) | 36.9 (36.5-37.2) | 0.319<sup>a</sup> |
| WBC ($\times 10^3/\mu\text{L}$), median [IQR] | 14.70 (11.90-17.13) | 14.05 (11.67-17.00) | 15.35 (12.07-17.30) | 0.327<sup>a</sup> |
| CRP (mg/L), median [IQR] | 49 (12-141) | 52 (14-118) | 45 (11-160) | 0.923<sup>a</sup> |
| **Imaging** |  |  |  |  |
| USG - Appendicitis confirmed, n (%) | 68 (31.6) | 38 (41.3) | 30 (24.4) | **0.013<sup>c\*</sup>** |
| USG - Not performed, n (%) | 96 (44.7) | 37 (40.2) | 59 (48.0) | — |
| CT - Appendicitis confirmed, n (%) | 133 (61.9) | 54 (58.7) | 79 (64.2) | 0.382<sup>c</sup> |
| CT - Not performed, n (%) | 53 (24.7) | 28 (30.4) | 25 (20.3) | — |
| **Surgeon Seniority** |  |  |  |  |
| Trainee surgeon, n (%) | 89 (41.4) | 39 (42.4) | 50 (40.7) | 0.798<sup>b</sup> |
| Consultant/attending surgeon, n (%) | 126 (58.6) | 53 (57.6) | 73 (59.3) | — |
| **Outcomes** |  |  |  |  |
| Unplanned readmission/reintervention, n (%) | 21 (9.8) | 6 (6.5) | 15 (12.2) | 0.166<sup>b</sup> |
| Clavien-Dindo IIIa (radiologic drainage) | 9 (4.2) | 3 (3.3) | 6 (4.9) | 0.411<sup>c</sup> |
| Clavien-Dindo IIIb (reoperation) | 12 (5.6) | 3 (3.3) | 9 (7.3) | — |

*AIR: Appendicitis Inflammatory Response score; AAS: Adult Appendicitis Score; CRP: C-reactive protein; IQR: interquartile range; LOS: length of stay; RIPASA: Raja Isteri Pengiran Anak Saleha Appendicitis score; USG: ultrasonography; WBC: white blood cell count. <sup>a</sup> Mann-Whitney U test; <sup>b</sup> Pearson's chi-square test; <sup>c</sup> Fisher's exact test. \* $p < 0.05$ .*

Baseline characteristics were well balanced between the daytime and night-time groups for age, sex, nationality, all appendicitis scoring systems, laboratory parameters, and surgeon seniority (all p>0.05). The only significant baseline difference was a higher rate of USG-confirmed appendicitis in the daytime group (41.3% vs. 24.4%, p=0.013). Weekend and weekday groups were balanced across all baseline variables. The overall median LOS was 2 days.

### Surgical timing analyses

Night-time surgery was associated with higher frequencies of unplanned readmission / reintervention (12.2% vs. 6.5%; OR 1.99 [95% CI 0.74-5.35], p=0.166) and conversion to open surgery (9.8% vs. 3.3%; OR 3.21 [0.88-11.72], p=0.064) compared with daytime surgery; however, neither comparison reached significance (Table 2). No significant effect of night-time surgery was observed for prolonged LOS (20.3% vs. 26.1%; OR 0.72 [0.38-1.37], p=0.319).

**Table 2.** Night vs. day surgery.

| Outcome | Day<br>(n=92) | Night<br>(n=123) | OR<br>[95% CI] | p-value |
| --- | --- | --- | --- | --- |
| Readmission/reintervention, n (%) | 6 (6.5) | 15 (12.2) | 1.99 [0.74-5.35] | 0.166<sup>a</sup> |
| Conversion to open surgery, n (%) | 3 (3.3) | 12 (9.8) | 3.21 [0.88-11.72] | 0.064<sup>a</sup> |
| Prolonged LOS >3 days, n (%) | 24 (26.1) | 25 (20.3) | 0.72 [0.38-1.37] | 0.319<sup>a</sup> |

<sup>a</sup> Pearson's chi-square test. LOS: length of stay; OR: odds ratio.

No significant weekend effect was observed for any outcome. Readmission/reintervention rates were comparable between weekend and weekday cases (7.0% vs. 10.5%; OR 0.64 [0.18-2.23], p=0.774), as were conversion rates (2.3% vs. 8.1%; OR 0.27 [0.03-2.10], p=0.314) and prolonged LOS (18.6% vs. 23.8%; OR 0.73 [0.31-1.70], p=0.464; Table 3).

**Table 3.** Weekend vs. weekday surgery.

| Outcome | Weekday<br>(n=172) | Weekend<br>(n=43) | OR<br>[95% CI] | p-value |
| --- | --- | --- | --- | --- |
| Readmission/reintervention, n (%) | 18 (10.5) | 3 (7.0) | 0.64 [0.18-2.23] | 0.774<sup>c</sup> |
| Conversion to open surgery, n (%) | 14 (8.1) | 1 (2.3) | 0.27 [0.03-2.10] | 0.314<sup>c</sup> |
| Prolonged LOS >3 days, n (%) | 41 (23.8) | 8 (18.6) | 0.73 [0.31-1.70] | 0.464<sup>a</sup> |

<sup>a</sup> Pearson's chi-square test; <sup>c</sup> Fisher's exact test. LOS: length of stay; OR: odds ratio.

### Surgeon seniority

Surgeon seniority was associated with the primary outcome (Table 4). Surgeries performed by trainees resulted in a significantly higher frequency of unplanned readmission / reintervention relative to consultants (15.7% vs. 5.6%; OR 0.32 [95% CI 0.12-0.82], p=0.013). Conversion rates were similar between groups (6.7% vs. 7.1%; OR 1.06 [0.37-3.10], p=0.909), as was prolonged LOS (19.1% vs. 25.4%; OR 1.44 [0.74-2.80], p=0.278). Trainees had performed 42.4% of daytime and 40.7% of night-time cases (p=0.798), confirming that seniority did not confound the timing analyses.

**Table 4.** Surgeon seniority.

| **Table 4.** Surgeon seniority |  |  |  |  |
| --- | --- | --- | --- | --- |
| **Outcome** | **Trainee**<br>**(n=89)** | **Consultant**<br>**(n=126)** | **OR**<br>**[95% CI]** | **p-value** |
| Readmission/reintervention†, n (%) | 14 (15.7) | 7 (5.6) | 0.32 [0.12-0.82] | **0.013<sup>a</sup> \*** |
| Conversion to open surgery, n (%) | 6 (6.7) | 9 (7.1) | 1.06 [0.37-3.10] | 0.909<sup>a</sup> |
| Prolonged LOS >3 days, n (%) | 17 (19.1) | 32 (25.4) | 1.44 [0.74-2.80] | 0.278<sup>a</sup> |
| *a* Pearson's chi-square test. LOS: length of stay; OR: odds ratio. * $p<0.05$. | | | | |

### Propensity score matching sensitivity analysis

PSM produced 88 matched pairs for the night vs. day comparison, 42 pairs for the weekend vs. weekday comparison, and 88 pairs for the trainee vs. consultant comparison (all SMD <0.1) (Table 5).

**Table 5.** Propensity score matching sensitivity analysis.

| Comparison / Outcome | Reference n (%) | Exposed n (%) | OR [95% CI] | p-value |
| --- | --- | --- | --- | --- |
| **PSM: Night vs. Day surgery - 88 matched pairs, SMD <0.1 (Reference: Day)** |  |  |  |  |
| Readmission/reintervention | 5 (5.7) | 12 (13.6) | 2.45 [0.85-7.03] | 0.09 |
| Conversion to open surgery | 3 (3.4) | 9 (10.2) | 2.84 [0.73-11.1] | 0.13 |
| Prolonged LOS >3 days | 24 (27.3) | 19 (21.6) | 0.79 [0.39-1.60] | 0.51 |
| **PSM: Weekend vs. Weekday surgery - 42 matched pairs, SMD <0.1 (Reference: Weekday)** |  |  |  |  |
| Readmission/reintervention | 2 (4.8) | 3 (7.1) | 1.53 [0.24-9.72] | 0.65 |
| Conversion to open surgery | 3 (7.1) | 1 (2.4) | 0.31 [0.03-3.17] | 0.32 |
| Prolonged LOS >3 days | 6 (14.3) | 8 (19.1) | 1.41 [0.44-4.52] | 0.55 |
| **PSM: Trainee vs. Consultant surgeon - 88 matched pairs, SMD <0.1 (Reference: Consultant)** |  |  |  |  |
| Readmission/reintervention | 4 (4.5) | 14 (15.9) | 0.25 [0.08-0.79] | **0.013\*** |
| Conversion to open surgery | 8 (9.1) | 6 (6.8) | 1.37 [0.45-4.12] | 0.577 |
| Prolonged LOS >3 days | 28 (31.8) | 17 (19.3) | 1.95 [0.97-3.90] | 0.057 |
| *PSM: propensity score matching; SMD: standardised mean difference; LOS: length of stay; OR: odds ratio.*<br>*Matching performed using RIPASA score as propensity variable; 1:1 nearest-neighbour matching without replacement;*<br>*caliper 0.05 on logit scale. \* $p < 0.05$.* |  |  |  |  |

In the matched night vs. day cohort, the numerically higher readmission/reintervention rate for night-time surgery persisted (OR 2.45 [0.85-7.03], p=0.09) and the conversion trend was maintained (OR 2.84 [0.73-11.1], p=0.13), both remaining non-significant. Weekend vs. weekday comparisons were likewise non-significant after matching (readmission OR 1.53 [0.24-9.72], p=0.65). The significant association between trainee seniority and readmission/reintervention was confirmed in the matched cohort (OR 0.25 [0.08-0.79], p=0.013), with a non-significant trend for prolonged LOS (OR 1.95 [0.97-3.90], p=0.057).

## Discussion

This propensity score-matched analysis of 215 patients undergoing laparoscopic appendectomy for histologically confirmed complicated appendicitis yielded two principal findings. First, surgical timing, whether defined by time of day or day of the week, was not associated with unplanned readmission, reintervention, or conversion to open surgery. Second, surgeon seniority was the only factor independently and significantly associated with the primary outcome in both primary and sensitivity analyses, with trainee-led surgery carrying a three-fold higher rate of unplanned readmission or reintervention compared with consultant-led procedures.

The absence of a significant timing effect aligns with the broader literature on appendectomy timing. A 2026 systematic review and meta-analysis by Hu et al. found that out-of-hours appendectomy was associated with a marginally higher conversion rate but no significant increase in postoperative complications, readmission, or mortality [7]. Similarly, a 2024 network meta-analysis by Calpin et al. found no significant effect of time-of-day surgery on readmission or complication rates in appendicectomy [8], and a meta-analysis across 33,596 patients found no significant night-time effect on overall complications, LOS, or readmission [11]. Our findings extend this evidence specifically to complicated appendicitis, a subgroup one might hypothesize to be more vulnerable to the consequences of suboptimal out-of-hours care given its greater intrinsic operative difficulty.

The non-significant trend toward higher conversion in the night-time group (9.8% vs. 3.3%, p=0.064; PSM-confirmed OR 2.84, p=0.13) warrants clinical attention despite not reaching statistical significance. The effect size is substantial, and the p-values in both the primary and matched analyses are consistent with underpowering rather than a true null effect. Night-time surgery may present additional challenges beyond the operative environment itself. Our data show that ultrasound-confirmed appendicitis was significantly less frequent at night (24.4% vs. 41.3%, p=0.013), whereas CT-confirmed appendicitis was numerically more frequent in the night-time group (64.2% vs. 58.7%, p=0.382). This suggests that out-of-hours diagnostic pathways may differ, with reduced reliance on ultrasonography and greater reliance on CT. Moreover, organizational factors, including reduced scrub nurse experience, limited intraoperative consultation, and differences in anaesthetic support, may compound surgical difficulty during out-of-hours procedures. These contextual factors are not captured in our dataset but deserve prospective investigation.

The absence of a weekend effect is consistent with findings from large administrative database studies. In a propensity score-matched analysis of 103,501 paediatric cases of complicated appendicitis, Lane et al. found no significant outcome differences between weekend and weekday laparoscopic appendectomy [18]. Similarly, Worni et al., in a national study of 151,774 patients undergoing laparoscopic appendectomy, reported no clinically significant weekend effect, with no significant differences in overall postoperative complications, mortality, or length of hospital stay after risk adjustment [19]. Our data corroborate these findings in an adult complicated appendicitis cohort, suggesting that weekend surgical care quality for this procedure is maintained at weekday standards, possibly because appendectomy is generally managed as an urgent surgical condition requiring timely operative care [20].

The finding that trainee surgeon status was independently and significantly associated with a three-fold higher rate of unplanned readmission and reintervention (15.7% vs. 5.6%, p=0.013, confirmed by PSM) is the most clinically impactful result of this study. It appears to contrast with the conclusion of the systematic review by Anyomih et al., who found that emergency appendectomy by trainees did not compromise patient safety [11]. However, that review was predominantly derived from mixed appendicitis populations, many of which included uncomplicated cases. Our analysis is restricted to complicated appendicitis, a setting characterized by greater tissue fragility, obscured anatomical landmarks, and a higher need for intraoperative decision-making. The principle of graded operative autonomy, described in the SnapAppy prospective study, holds that case complexity should drive attending involvement; our data suggest that in complicated appendicitis specifically, this principle may not be consistently applied with sufficient rigor [10].

Crucially, the absence of a confounding relationship between seniority and timing in our cohort (trainee rate: 42.4% day vs. 40.7% night, p=0.798) indicates that the seniority-outcome association is not attributable to trainees being disproportionately allocated to simpler daytime cases. This strengthens the causal plausibility of the finding and suggests that the issue is not delegation per se, but rather the adequacy of senior oversight during trainee-led procedures, particularly in the out-of-hours context, where consultant availability may be limited.

### Limitations

Several limitations merit acknowledgement. First, the retrospective secondary analysis precludes the analysis of individual surgeon experience, institutional volume, or intraoperative details, which might have had marginal effects on the analyses. It is notable that some non-significant findings, especially the night-time conversion trend, may reflect underpowered statistical results; however, it was necessary to adhere to the strict exclusion criteria in order to arrive upon reliable conclusions for the examined patient subset. Third, the RIFT Turkey data were collected during the early COVID-19 pandemic (September-December 2020), a period of organizational disruption and restricted elective activity, potentially limiting generalizability to the current era. Fourth, although seniority was prospectively recorded, the differences between centers in terms of supervision intensity, intraoperative attending involvement, or individual case volume were unavailable, limiting the interpretations in this regard.

### Conclusions

In this propensity score-matched analysis of complicated appendicitis patients from the RIFT Turkey cohort, surgical timing, whether night versus day or weekend versus weekday, was not significantly associated with short-term postoperative outcomes. Night-time surgery showed a consistent, clinically meaningful trend towards higher conversion rates that did not reach statistical significance, likely reflecting insufficient power. Surgeon seniority was the only factor independently and significantly associated with unplanned readmission and reintervention in both primary and matched analyses. These findings support the prioritization of adequate consultant oversight in the management of complicated appendicitis, and suggest that supervision standards, not simply surgical scheduling, are the modifiable determinant of outcomes in this high-risk population. Prospective studies with larger cohorts are needed to characterize the interaction between timing, surgeon experience, and patient outcomes in complicated appendicitis.

## Supporting information

Authorship appendix

## Data Availability

Data are available from the corresponding author upon reasonable request.

## Declarations

### Ethics approval and consent to participate

Ethical approval was obtained from the Clinical Research Ethics Committee of Gazi University Faculty of Medicine (7 September 2020). The study was registered on ClinicalTrials.gov (NCT04614649). Written informed consent was obtained from all participants in the parent RIFT Turkey study.

### Availability of data and materials

Data are available from the corresponding author upon reasonable request.

### Authors’ contributions

AY and SDA contributed equally and share first authorship. SDA, CO, AY, and AhY conceived the study design. CO, DK, AY, AhY performed the statistical analysis. AY, CO, and SDA wrote the first draft. EK, AhY, and SL critically revised the manuscript for important intellectual content. All authors approved the final version.

### Collaborators

Ali Yalcinkaya (Gazi University Hospital, Ankara, Turkey); Ahmet Yalcinkaya (Uppsala University, Uppsala, Sweden); Can Keskin (Tibbi Akademik, Ankara, Turkey); Ibrahim Erkan (Tibbi Akademik, Ankara, Turkey); Sezai Leventoglu (Gazi University Hospital, Ankara, Turkey); Mehmet Caglikulekci (Istanbul Yeni Yuzyıl University Gaziosmanpasa Hospital, Istanbul, Turkey); Elbrus Zarbaliyev (Istanbul Yeni Yuzyıl University Gaziosmanpasa Hospital, Istanbul, Turkey); Murat Sevmis (Istanbul Yeni Yuzyıl University Gaziosmanpasa Hospital, Istanbul, Turkey); Yigit Ulgen (Bagcilar Training and Research Hospital, Istanbul, Turkey); Yuksel Altinel (Bagcilar Training and Research Hospital, Istanbul, Turkey); Serhat Meric (Bagcilar Training and Research Hospital, Istanbul, Turkey); Ahmet Akbas (Bagcilar Training and Research Hospital, Istanbul, Turkey); Nadir Adnan Hacim (Bagcilar Training and Research Hospital, Istanbul, Turkey); Talar Vartanoglu Aktokmanyan (Bagcilar Training and Research Hospital, Istanbul, Turkey); Yunus Emre Aktimur (Bagcilar Training and Research Hospital, Istanbul, Turkey); Fikret Calikoglu (Bagcilar Training and Research Hospital, Istanbul, Turkey); Hasim Furkan Gullu (Bagcilar Training and Research Hospital, Istanbul, Turkey); Ahmet Guray Durma (Bagcilar Training and Research Hospital, Istanbul, Turkey); Sami Acar (Zeynep Kamil Women’s and Children’s Diseases Training and Research Hospital, Istanbul, Turkey); Erman Ciftci (Zeynep Kamil Women’s and Children’s Diseases Training and Research Hospital, Istanbul, Turkey); Emre Balik (Koc University Hospital, Istanbul, Turkey); Cemil Burak Kulle (Koc University Hospital, Istanbul, Turkey); Ibrahim Halil Ozata (Koc University Hospital, Istanbul, Turkey); Tutku Tufekci (Koc University Hospital, Istanbul, Turkey); Cihad Tatar (Istanbul Training and Research Hospital, Istanbul, Turkey); Mert Mahsuni Sevinc (Istanbul Training and Research Hospital, Istanbul, Turkey); Husnu Sevik (Istanbul Training and Research Hospital, Istanbul, Turkey); Candeniz Ertürk (Istanbul Training and Research Hospital, Istanbul, Turkey); Irem Nur Kiraz (Istanbul Training and Research Hospital, Istanbul, Turkey); Volkan Ozben (Acibadem Mehmet Ali Aydinlar University Atakent Hospital, Istanbul, Turkey); Erman Aytac (Acibadem Mehmet Ali Aydinlar University Atakent Hospital, Istanbul, Turkey); Zumrud Aliyeva (Acibadem Mehmet Ali Aydinlar University Atakent Hospital, Istanbul, Turkey); Arda Ulas Mutlu (Acibadem Mehmet Ali Aydinlar University Atakent Hospital, Istanbul, Turkey); Mert Tanal (University of Health Sciences, Sisli Hamidiye Etfal Training and Research Hospital, Istanbul, Turkey); Mustafa Fevzi Celayir (University of Health Sciences, Sisli Hamidiye Etfal Training and Research Hospital, Istanbul, Turkey); Emre Bozkurt (University of Health Sciences, Sisli Hamidiye Etfal Training and Research Hospital, Istanbul, Turkey); Sitki Gurkan Yetkin (University of Health Sciences, Sisli Hamidiye Etfal Training and Research Hospital, Istanbul, Turkey); Emin Ergin (University of Health Sciences, Sisli Hamidiye Etfal Training and Research Hospital, Istanbul, Turkey); Wafi Attaallah (Marmara University Hospital, Istanbul, Turkey); Tevfik Kivilcim Uprak (Marmara University Hospital, Istanbul, Turkey); Ahmet Omak (Marmara University Hospital, Istanbul, Turkey); Oguzhan Simsek (Marmara University Hospital, Istanbul, Turkey); Mehmet Abdussamet Bozkurt (Kanuni Sultan Suleyman Training and Research Hospital, Istanbul, Turkey); Yasin Kara (Kanuni Sultan Suleyman Training and Research Hospital, Istanbul, Turkey); Emre Bozdag (Kanuni Sultan Suleyman Training and Research Hospital, Istanbul, Turkey); Hakan Yirgin (Kanuni Sultan Suleyman Training and Research Hospital, Istanbul, Turkey); Adem Ozcan (Kanuni Sultan Suleyman Training and Research Hospital, Istanbul, Turkey); Nuri Okkabaz (Medipol University Medipol Mega Hospital, Istanbul, Turkey); Yasar Ozdenkaya (Medipol University Medipol Mega Hospital, Istanbul, Turkey); Mustafa Celalettin Haksal (Medipol University Medipol Mega Hospital, Istanbul, Turkey); Caglar Kazim Pekuz (Medipol University Medipol Mega Hospital, Istanbul, Turkey); Sila Duru (Medipol University Medipol Mega Hospital, Istanbul, Turkey); Emre Sivrikoz (Acıbadem Bakirkoy Hospital, Istanbul, Turkey); Yavuz Ozdemir (Pendik Yuzyil Private Hospital, Istanbul, Turkey); Necati Tan (Pendik Yuzyil Private Hospital, Istanbul, Turkey); Feza Yarbug Karayali (Baskent University Istanbul Hospital, Istanbul, Turkey); Abdulla Taghiyeva (Baskent University Istanbul Hospital, Istanbul, Turkey); Ismail Tirnova (Baskent University Istanbul Hospital, Istanbul, Turkey); Ilknur Erenler Bayraktar (Istanbul Florence Nightingale Hospital, Istanbul, Turkey); Onur Bayraktar (Istanbul Florence Nightingale Hospital, Istanbul, Turkey); Emine Zulal Emsal (Istanbul Florence Nightingale Hospital, Istanbul, Turkey); Munevver Irem Dalkilic (Istanbul Florence Nightingale Hospital, Istanbul, Turkey); Metin Yesiltas (Professor Cemil Tascioglu City Hospital, Istanbul, Turkey); Hasan Tok (Professor Cemil Tascioglu City Hospital, Istanbul, Turkey); Dursun Ozgur Karakas (Professor Cemil Tascioglu City Hospital, Istanbul, Turkey); Ali Pusane (Professor Cemil Tascioglu City Hospital, Istanbul, Turkey); Ali Ilbey Demirer (Professor Cemil Tascioglu City Hospital, Istanbul, Turkey); Hasan Berk Sahin (Professor Cemil Tascioglu City Hospital, Istanbul, Turkey); Ali Fuat Kaan Gok (Istanbul University, Istanbul Faculty of Medicine, Istanbul, Turkey); Halil Alper Bozkurt (Istanbul University, Istanbul Faculty of Medicine, Istanbul, Turkey); Mehmet Iskender Yildirim (Istanbul University, Istanbul Faculty of Medicine, Istanbul, Turkey); Gorkem Uzunyolcu (Istanbul University, Istanbul Faculty of Medicine, Istanbul, Turkey); Hakan Teoman Yanar (Istanbul University, Istanbul Faculty of Medicine, Istanbul, Turkey); Sefa Ergun (Istanbul University-Cerrahpasa, Cerrahpasa Faculty of Medicine, Istanbul, Turkey); Fadime Kutluk (Istanbul University-Cerrahpasa, Cerrahpasa Faculty of Medicine, Istanbul, Turkey); Server Sezgin Uludag (Istanbul University-Cerrahpasa, Cerrahpasa Faculty of Medicine, Istanbul, Turkey); Abdullah Kagan Zengin (Istanbul University-Cerrahpasa, Cerrahpasa Faculty of Medicine, Istanbul, Turkey); Mehmet Faik Ozcelik (Istanbul University-Cerrahpasa, Cerrahpasa Faculty of Medicine, Istanbul, Turkey); Ahmet Necati Sanli (Istanbul University-Cerrahpasa, Cerrahpasa Faculty of Medicine, Istanbul, Turkey); Yunus Emre Altuntas (Kartal Dr. Lutfi Kirdar City Hospital, Istanbul, Turkey); Ecem Memisoglu (Kartal Dr. Lutfi Kirdar City Hospital, Istanbul, Turkey); Ramazan Sari (Kartal Dr. Lutfi Kirdar City Hospital, Istanbul, Turkey); Osman Akdogan (Kartal Dr. Lutfi Kirdar City Hospital, Istanbul, Turkey); Hasan Fehmi Kucuk (Kartal Dr. Lutfi Kirdar City Hospital, Istanbul, Turkey); Omer Faruk Ozkan (Istanbul Umraniye Training and Research Hospital, Istanbul, Turkey); Hanife Seyda Ulgur (Istanbul Umraniye Training and Research Hospital, Istanbul, Turkey); Emre Furkan Kirkan (Istanbul Umraniye Training and Research Hospital, Istanbul, Turkey); Sema Yuksekdag (Istanbul Umraniye Training and Research Hospital, Istanbul, Turkey); Ahmet Rencuzogullari (Cukurova University Hospital, Adana, Turkey); Melik Kagan Aktas (Cukurova University Hospital, Adana, Turkey); Murat Aba (Cukurova University Hospital, Adana, Turkey); Ahmet Onur Demirel (Cukurova University Hospital, Adana, Turkey); Ismail Cem Eray (Cukurova University Hospital, Adana, Turkey); Burak Aydogan (Cukurova University Hospital, Adana, Turkey); Suleyman Cetinkunar (University of Health Sciences, Adana City Training and Research Hospital, Adana, Turkey); Kemal Yener (University of Health Sciences, Adana City Training and Research Hospital, Adana, Turkey); Alper Sozutek (University of Health Sciences, Adana City Training and Research Hospital, Adana, Turkey); Oktay Irkorucu (University of Health Sciences, Adana City Training and Research Hospital, Adana, Turkey); Mehmet Bayrak (Adana Ortadogu Private Hospital, Adana, Turkey); Yasemin Altintas (Adana Ortadogu Private Hospital, Adana, Turkey); Omer Alabaz (Adana Ortadogu Private Hospital, Adana, Turkey); Ahmet Atasever (Kahramanmaras Afsin District State Hospital, Kahramanmaras, Turkey); Guven Erdogrul (Kahramanmaras Afsin District State Hospital, Kahramanmaras, Turkey); Aydın Hakan Kupeli (Necip Fazil City Hospital, Kahramanmaras, Turkey); Bahtiyar Muhammedoglu (Necip Fazil City Hospital, Kahramanmaras, Turkey); Suleyman Kokdas (Necip Fazil City Hospital, Kahramanmaras, Turkey); Murat Kaya (Necip Fazil City Hospital, Kahramanmaras, Turkey); Erkan Uysal (Necip Fazil City Hospital, Kahramanmaras, Turkey); Ali Cihat Yildirim (University of Health Sciences, Kutahya Evliya Celebi Training and Research Hospital, Kutahya, Turkey); Sezgin Zeren (University of Health Sciences, Kutahya Evliya Celebi Training and Research Hospital, Kutahya, Turkey); Mehmet Fatih Ekici (University of Health Sciences, Kutahya Evliya Celebi Training and Research Hospital, Kutahya, Turkey); Mustafa Cem Algin (University of Health Sciences, Kutahya Evliya Celebi Training and Research Hospital, Kutahya, Turkey); Gultekin Ozan Kucuk (University of Health Sciences, Samsun Training and Research Hospital, Samsun, Turkey); Huseyin Eraslan (University of Health Sciences, Samsun Training and Research Hospital, Samsun, Turkey); Engin Aybar (University of Health Sciences, Samsun Training and Research Hospital, Samsun, Turkey); Suleyman Polat (University of Health Sciences, Samsun Training and Research Hospital, Samsun, Turkey); Alper Ceylan (University of Health Sciences, Samsun Training and Research Hospital, Samsun, Turkey); Ozgen Isik (Uludag University Hospital, Bursa, Turkey); Said Kural (Uludag University Hospital, Bursa, Turkey); Ahmet Aktas (Uludag University Hospital, Bursa, Turkey); Burak Bakar (Uludag University Hospital, Bursa, Turkey); Mustafa Yener Uzunoglu (Kestel State Hospital, Bursa, Turkey); Baris Gulcu (Medicana Bursa Hospital, Bursa, Turkey); Ersin Ozturk (Medicana Bursa Hospital, Bursa, Turkey); Ali Onder Devay (Medicana Bursa Hospital, Bursa, Turkey); Ersoy Taspinar (Medicana Bursa Hospital, Bursa, Turkey); Ozkan Balcin (Bursa City Hospital, Bursa, Turkey); Fuat Aksoy (Bursa City Hospital, Bursa, Turkey); Gokhan Garip (Bursa City Hospital, Bursa, Turkey); Omer Yalkin (Bursa City Hospital, Bursa, Turkey); Nidal Iflazoglu (Bursa City Hospital, Bursa, Turkey); Direnc Yigit (Bursa City Hospital, Bursa, Turkey); Rumeysa Betul Kaya (Bursa City Hospital, Bursa, Turkey); Mustafa Ugur (Hatay Mustafa Kemal University Hospital, Hatay, Turkey); Erol Kilic (Hatay Mustafa Kemal University Hospital, Hatay, Turkey); Akin Dedemoglu (Hatay Mustafa Kemal University Hospital, Hatay, Turkey); Rasim Ersin Arslan (Hatay Mustafa Kemal University Hospital, Hatay, Turkey); Muhyittin Temiz (Hatay Mustafa Kemal University Hospital, Hatay, Turkey); Cengiz Aydin (University of Health Sciences, Tepecik Training and Research Hospital, Izmir, Turkey); Semra Demirli Atici (University of Health Sciences, Tepecik Training and Research Hospital, Izmir, Turkey); Tayfun Kaya (University of Health Sciences, Tepecik Training and Research Hospital, Izmir, Turkey); Selen Ozturk (University of Health Sciences, Tepecik Training and Research Hospital, Izmir, Turkey); Bulent Calik (University of Health Sciences, Tepecik Training and Research Hospital, Izmir, Turkey); Gizem Kilinc (University of Health Sciences, Tepecik Training and Research Hospital, Izmir, Turkey); Erdinc Kamer (Izmir Katip Celebi University Ataturk Training and Research Hospital, Izmir, Turkey); Turan Acar (Izmir Katip Celebi University Ataturk Training and Research Hospital, Izmir, Turkey); Nihan Acar (Izmir Katip Celebi University Ataturk Training and Research Hospital, Izmir, Turkey); Fevzi Cengiz (Izmir Katip Celebi University Ataturk Training and Research Hospital, Izmir, Turkey); Orhan Ureyen (University of Health Sciences, Izmir Bozyaka Training and Research Hospital, Izmir, Turkey); Sedat Tan (University of Health Sciences, Izmir Bozyaka Training and Research Hospital, Izmir, Turkey); Mehmet Yildirim (University of Health Sciences, Izmir Bozyaka Training and Research Hospital, Izmir, Turkey); Enver Ilhan (University of Health Sciences, Izmir Bozyaka Training and Research Hospital, Izmir, Turkey); Yigit Turk (Bakırçay University Cigli Training and Research Hospital, Izmir, Turkey); Ahmet Turan Durak (Urla State Hospital, Izmir, Turkey); Mehmet Yilmaz (Buca Seyfi Demirsoy Training and Research Hospital, Izmir, Turkey); Metin Mercan (Buca Seyfi Demirsoy Training and Research Hospital, Izmir, Turkey); Recep Atci (Buca Seyfi Demirsoy Training and Research Hospital, Izmir, Turkey); Selman Sokmen (Dokuz Eylul University Hospital, Izmir, Turkey); Tayfun Bisgin (Dokuz Eylul University Hospital, Izmir, Turkey); Tufan Egeli (Dokuz Eylul University Hospital, Izmir, Turkey); Yasemin Yildirim (Dokuz Eylul University Hospital, Izmir, Turkey); Turugsan Safak (Dokuz Eylul University Hospital, Izmir, Turkey); Kazim Celik (Tire State Hospital, Izmir, Turkey); Eyup Murat Yilmaz (Aydın Adnan Menderes University Hospital, Aydin, Turkey); Mahir Kirnap (Aydın Adnan Menderes University Hospital, Aydin, Turkey); Ahmet Ender Demirkiran (Aydın Adnan Menderes University Hospital, Aydin, Turkey); Ulas Utku Sekerci (Aydın Adnan Menderes University Hospital, Aydin, Turkey); Erkan Karacan (Aydin State Hospital, Aydin, Turkey); Ethem Bilgic (Didim State Hospital, Aydin, Turkey); Mehmet Mahir Ozmen (Liv Hospital Ankara, Ankara, Turkey); Cem Emir Guldogan (Liv Hospital Ankara, Ankara, Turkey); Emre Gundogdu (Liv Hospital Ankara, Ankara, Turkey); Munevver Moran (Liv Hospital Ankara, Ankara, Turkey); Timucin Erol (Hacettepe University Hospital, Ankara, Turkey); Hilmi Anil Dincer (Hacettepe University Hospital, Ankara, Turkey); Busenur Kirimtay (Hacettepe University Hospital, Ankara, Turkey); Sumeyye Yilmaz (Hacettepe University Hospital, Ankara, Turkey); Omer Cennet (Hacettepe University Hospital, Ankara, Turkey); Alp Yildiz (Yildirim Beyazit University Yenimahalle Training and Research Hospital, Ankara, Turkey); Aybala Yildiz (Yildirim Beyazit University Yenimahalle Training and Research Hospital, Ankara, Turkey); Can Sahin (Yildirim Beyazit University Yenimahalle Training and Research Hospital, Ankara, Turkey); Cihangir Akyol (Ankara University Ibni Sina Hospital, Ankara, Turkey); Mehmet Ali Koc (Ankara University Ibni Sina Hospital, Ankara, Turkey); Siyar Ersoz (Ankara University Ibni Sina Hospital, Ankara, Turkey); Anil Turhan (Ankara University Ibni Sina Hospital, Ankara, Turkey); Can Konca (Ankara University Ibni Sina Hospital, Ankara, Turkey); Tugan Tezcaner (Baskent University Ankara Hospital, Ankara, Turkey); Murathan Erkent (Baskent University Ankara Hospital, Ankara, Turkey); Onur Aydin (Baskent University Ankara Hospital, Ankara, Turkey); Tevfik Avci (Baskent University Ankara Hospital, Ankara, Turkey); Saygin Altiner (Ankara Training and Research Hospital, Ankara, Turkey); Igbal Osmanov (Memorial Ankara Hospital, Ankara, Turkey); Ahmet Cihangir Emral (Sincan State Hospital, Ankara, Turkey); Gokay Cetinkaya (Sincan State Hospital, Ankara, Turkey); Emin Lapsekili (Gulhane Training and Research Hospital, Ankara, Turkey); Merve Sakca (Gulhane Training and Research Hospital, Ankara, Turkey); Sebnem Cimen (Gulhane Training and Research Hospital, Ankara, Turkey); Dogan Ozen (Gulhane Training and Research Hospital, Ankara, Turkey); Erdem Baran Kozan (Gulhane Training and Research Hospital, Ankara, Turkey); Lutfi Dogan (Ankara Oncology Training and Research Hospital, Ankara, Turkey); Elifcan Haberal (Ankara Oncology Training and Research Hospital, Ankara, Turkey); Bengi Balci (Ankara Oncology Training and Research Hospital, Ankara, Turkey); Okan Kayhan (Ankara Oncology Training and Research Hospital, Ankara, Turkey); Bulent Aksel (Ankara Oncology Training and Research Hospital, Ankara, Turkey); Harun Karabacak (University of Health Sciences, Ankara Diskapi Yildirim Beyazid Training and Research Hospital, Ankara, Turkey); Cem Azili (University of Health Sciences, Ankara Diskapi Yildirim Beyazid Training and Research Hospital, Ankara, Turkey); Faruk Yazici (University of Health Sciences, Ankara Diskapi Yildirim Beyazid Training and Research Hospital, Ankara, Turkey); Muhammed Apaydin (University of Health Sciences, Ankara Diskapi Yildirim Beyazid Training and Research Hospital, Ankara, Turkey); Ismail Oskay Kaya (University of Health Sciences, Ankara Diskapi Yildirim Beyazid Training and Research Hospital, Ankara, Turkey); Erdinc Cetinkaya (Ankara City Hospital, Ankara, Turkey); Tezcan Akin (Ankara City Hospital, Ankara, Turkey); Gizem Gunes (Ankara City Hospital, Ankara, Turkey); Huseyin Turap (Ankara City Hospital, Ankara, Turkey); Deniz Aslan (Ankara City Hospital, Ankara, Turkey); Ali Eba Demirbag (Ankara City Hospital, Ankara, Turkey); Basak Bolukbasi (Gazi University Hospital, Ankara, Turkey); Berkay Enes Karaca (Gazi University Hospital, Ankara, Turkey); Ece Ozturk (Gazi University Hospital, Ankara, Turkey); Elif Ozeller (Gazi University Hospital, Ankara, Turkey); Gulsum Sueda Kayacan (Gazi University Hospital, Ankara, Turkey); Alp Ozgun Borcek (Gazi University Hospital, Ankara, Turkey); Ilhan Ece (Selcuk University Hospital, Konya, Turkey); Serdar Yormaz (Selcuk University Hospital, Konya, Turkey); Bayram Colak (Selcuk University Hospital, Konya, Turkey); Akin Calisir (Selcuk University Hospital, Konya, Turkey); Mustafa Sahin (Selcuk University Hospital, Konya, Turkey); Kemal Arslan (Konya City Hospital, Konya, Turkey); Ismail Hasirci (Konya City Hospital, Konya, Turkey); Mehmet Esref Ulutas (Konya City Hospital, Konya, Turkey); Sukru Hakan Metin (Konya City Hospital, Konya, Turkey); Fatma Ayca Gultekin (Zonguldak Bulent Ecevit University Hospital, Zonguldak, Turkey); Zeynep Ozkan (Elazig Fethi Sekin City Hospital, Elazig, Turkey); Onur Ilhan (Elazig Fethi Sekin City Hospital, Elazig, Turkey); Tamer Gundogdu (Elazig Fethi Sekin City Hospital, Elazig, Turkey); Rumeysa Kevser Liman (Elazig Fethi Sekin City Hospital, Elazig, Turkey); Burhan Hakan Kanat (Elazig Fethi Sekin City Hospital, Elazig, Turkey); Altan Aydin (University of Health Sciences, Trabzon Kanuni Training and Research Hospital, Trabzon, Turkey); Ugur Sungurtekin (Pamukkale University Hospital, Denizli, Turkey); Utku Ozgen (Pamukkale University Hospital, Denizli, Turkey); Muhammed Rasid Aykota (Pamukkale University Hospital, Denizli, Turkey); Fatih Altintoprak (Sakarya University Training and Research Hospital, Sakarya, Turkey); Emre Gonullu (Sakarya University Training and Research Hospital, Sakarya, Turkey); Guner Cakmak (Sakarya University Training and Research Hospital, Sakarya, Turkey); Ugur Can Dulger (Sakarya University Training and Research Hospital, Sakarya, Turkey); Baris Mantoglu (Sakarya University Training and Research Hospital, Sakarya, Turkey); Hakan Demir (Sakarya University Training and Research Hospital, Sakarya, Turkey); Emrah Akin (Sakarya University Training and Research Hospital, Sakarya, Turkey); Erhan Eroz (Toyotasa Emergency Hospital, Sakarya, Turkey); Okay Nazli (Mugla Sitki Kocman University Training and Research Hospital, Mugla, Turkey); Ozcan Dere (Mugla Sitki Kocman University Training and Research Hospital, Mugla, Turkey); Mustafa Aykut Dadasoglu (Mugla Sitki Kocman University Training and Research Hospital, Mugla, Turkey); Eray Kara (Manisa Celal Bayar University Hospital, Manisa, Turkey); Semra Tutcu (Manisa Celal Bayar University Hospital, Manisa, Turkey); Ilhami Solak (Manisa Celal Bayar University Hospital, Manisa, Turkey); Ilayda Gencer (Manisa Celal Bayar University Hospital, Manisa, Turkey); Alperen Dalkiran (Manisa Celal Bayar University Hospital, Manisa, Turkey); Baris Sevinc (Usak Training and Research Hospital, Usak, Turkey); Omer Karahan (Usak Training and Research Hospital, Usak, Turkey); Nurullah Damburaci (Usak Training and Research Hospital, Usak, Turkey); Erdem Sari (Bandirma State Hospital, Balikesir, Turkey); Tamer Akay (Bandirma State Hospital, Balikesir, Turkey); Alpaslan Fedayi Calta (Bandirma State Hospital, Balikesir, Turkey); Abdullah Ozdemir (Bandirma State Hospital, Balikesir, Turkey); Nurian Ohri (Balikesir State Hospital, Balikesir, Turkey); Ilker Ermis (Kirikkale Yuksek Ihtisas Hospital, Kirikkale, Turkey); Osman Bozbiyik (Ege University Hospital, Izmir, Turkey); Murat Ozdemir (Ege University Hospital, Izmir, Turkey); Berk Goktepe (Ege University Hospital, Izmir, Turkey); Batuhan Demir (Ege University Hospital, Izmir, Turkey); Ozgur Kilincarslan (Ege University Hospital, Izmir, Turkey); Umut Riza Gunduz (Antalya Training and Research Hospital, Antalya, Turkey); Mehmet Olcum (Antalya Training and Research Hospital, Antalya, Turkey); Onur Ilkay Dincer (Antalya Training and Research Hospital, Antalya, Turkey); Remzi Can Cakir (Antalya Training and Research Hospital, Antalya, Turkey); Bulent Dinc (Antalya Training and Research Hospital, Antalya, Turkey); Enes Sahin (Kocaeli State Hospital, Kocaeli, Turkey); Emrah Uludag (Kocaeli State Hospital, Kocaeli, Turkey); Yusuf Arslan (Kocaeli State Hospital, Kocaeli, Turkey); Gokhan Posteki (Kocaeli State Hospital, Kocaeli, Turkey); Ahmet Oktay (Kocaeli State Hospital, Kocaeli, Turkey); Ozan Can Tatar (Kocaeli University Hospital, Kocaeli, Turkey); Sertac Ata Guler (Kocaeli University Hospital, Kocaeli, Turkey); Nihat Zafer Utkan (Kocaeli University Hospital, Kocaeli, Turkey); Serkan Tayar (Erzurum Regional Training and Research Hospital, Erzurum, Turkey); Yasar Copelci (Erzurum Regional Training and Research Hospital, Erzurum, Turkey); Murat Kartal (Erzurum Regional Training and Research Hospital, Erzurum, Turkey); Tolga Kalayci (Erzurum Regional Training and Research Hospital, Erzurum, Turkey); Mustafa Yeni (Erzurum Regional Training and Research Hospital, Erzurum, Turkey); Ahmet Cagri Buyukkasap (Siirt State Hospital, Siirt, Turkey); Selahattin Vural (Giresun University Faculty of Medicine Training and Research Hospital, Giresun, Turkey); Tugrul Kesicioglu (Giresun University Faculty of Medicine Training and Research Hospital, Giresun, Turkey); Ismail Aydin (Giresun University Faculty of Medicine Training and Research Hospital, Giresun, Turkey); Mehmet Gulmez (Giresun University Faculty of Medicine Training and Research Hospital, Giresun, Turkey); Can Saracoglu (Giresun University Faculty of Medicine Training and Research Hospital, Giresun, Turkey); Omer Topcu (Sivas Cumhuriyet University Hospital, Sivas, Turkey); Atilla Kurt (Sivas Cumhuriyet University Hospital, Sivas, Turkey); Sinan Soylu (Sivas Cumhuriyet University Hospital, Sivas, Turkey); Begum Kurt (Sivas Cumhuriyet University Hospital, Sivas, Turkey); Musa Serin (Sivas Cumhuriyet University Hospital, Sivas, Turkey); Salim Ilksen Basceken (Diyarbakir Gazi Yasargil Training and Research Hospital, Diyarbakir, Turkey); Ebubekir Gundes (Diyarbakir Gazi Yasargil Training and Research Hospital, Diyarbakir, Turkey); Mervan Savda (Diyarbakir Gazi Yasargil Training and Research Hospital, Diyarbakir, Turkey); Ali Zeynel Abidin Balkan (Diyarbakir Gazi Yasargil Training and Research Hospital, Diyarbakir, Turkey); Mehmet Nuri Yildiz (Diyarbakir Gazi Yasargil Training and Research Hospital, Diyarbakir, Turkey); Ali Uzunkoy (Harran University Training and Research Hospital, Sanliurfa, Turkey); Emre Karaca (Harran University Training and Research Hospital, Sanliurfa, Turkey); Ahmet Berkan (Harran University Training and Research Hospital, Sanliurfa, Turkey); Arda Isik (Erzincan University Hospital, Erzincan, Turkey); Yasin Alper Yildiz (Kastamonu Training and Research Hospital, Kastamonu, Turkey); Zafer Ergul (Kastamonu Training and Research Hospital, Kastamonu, Turkey); Necdet Fatih Yasar (Eskisehir Osmangazi University Hospital, Eskisehir, Turkey); Bartu Badak (Eskisehir Osmangazi University Hospital, Eskisehir, Turkey); Ata Ozen (Eskisehir Osmangazi University Hospital, Eskisehir, Turkey); Melih Velipasaoglu (Eskisehir Osmangazi University Hospital, Eskisehir, Turkey); Iyimser Ure (Eskisehir Osmangazi University Hospital, Eskisehir, Turkey).

