## Supplementary material for "Surgical outcomes in complicated appendicitis: does timing or surgeon seniority matter? A propensity score-matched analysis from the RIFT Turkey cohort": Authorship appendix

**Appendix: Authorship (all co-authors are PubMed-indexed)**

**Collaborators (investigation, data collection, coordination, patient involvement):**

**(All collaborators are written in random order)**

1. Istanbul Yeni Yuzyıl University Gaziosmanpasa Hospital: Mehmet Caglikulekci, Elbrus Zarbaliyev, Murat Sevmis
2. Bagcilar Training and Research Hospital-1: Yigit Ulgen, Yuksel Altinel, Serhat Meric, Ahmet Akbas, Nadir Adnan Hacim
3. Bagcilar Training and Research Hospital-2: Talar Vartanoglu Aktokmanyan, Yunus Emre Aktimur, Fikret Calikoglu, Hasim Furkan Gullu, Ahmet Guray Durma
4. Zeynep Kamil Women’s and Children's Diseases Training and Research Hospital: Sami Acar, Erman Ciftci
5. Koc University Hospital: Emre Balik, Cemil Burak Kulle, Ibrahim Halil Ozata, Tutku Tufekci
6. Istanbul Training and Research Hospital: Cihad Tatar, Mert Mahsuni Sevinc, Husnu Sevik, Candeniz Ertürk, Irem Nur Kiraz
7. Acibadem Mehmet Ali Aydinlar University Atakent Hospital: Volkan Ozben, Erman Aytac, Zumrud Aliyeva, Arda Ulas Mutlu
8. University of Health Sciences, Sisli Hamidiye Etfal Training and Research Hospital: Mert Tanal, Mustafa Fevzi Celayir, Emre Bozkurt, Sitki Gurkan	Yetkin, Emin Ergin
9. Marmara University Hospital: Wafi Attaallah, Tevfik Kivilcim Uprak, Ahmet Omak, Oguzhan Simsek
10. Kanuni Sultan Suleyman Training and Research Hospital: Mehmet Abdussamet Bozkurt, Yasin Kara, Emre Bozdağ, Hakan Yirgin, Adem Ozcan
11. Medipol University Medipol Mega Hospital: Nuri Okkabaz, Yasar Ozdenkaya, Mustafa Celalettin Haksal, Caglar Kazim Pekuz, Sila Duru
12. Acıbadem Bakirkoy Hospital: Emre Sivrikoz
13. Pendik Yuzyil Private Hospital: Yavuz Ozdemir, Necati Tan
14. Baskent University Istanbul Hospital: Feza Yarbug Karayali, Abdulla Taghiyeva, Ismail Tirnova
15. Istanbul Florence Nightingale Hospital: Ilknur Erenler Bayraktar, Onur Bayraktar, Emine Zulal Emsal, Munevver Irem Dalkilic
16. Professor Cemil Tascioglu City Hospital: Metin Yesiltas, Hasan Tok, Dursun Ozgur Karakas, Ali Pusane, Ali Ilbey Demirer, Hasan Berk Sahin
17. Istanbul University, Istanbul Faculty of Medicine: Ali Fuat Kaan Gok, Halil Alper Bozkurt, Mehmet Iskender Yildirim, Gorkem Uzunyolcu, Hakan Teoman Yanar
18. Istanbul University-Cerrahpasa, Cerrahpasa Faculty of Medicine: Sefa Ergun, Fadime Kutluk, Server Sezgin Uludag, Abdullah Kagan Zengin, Mehmet Faik Ozcelik, Ahmet Necati Sanli
19. Kartal Dr. Lutfi Kirdar City Hospital: Yunus Emre Altuntas, Ecem Memisoglu, Ramazan Sari, Osman Akdogan, Hasan Fehmi Kucuk
20. Istanbul Umraniye Training and Research Hospital: Omer Faruk Ozkan, Hanife Seyda Ulgur, Emre Furkan Kirkan, Sema Yuksekdag
21. Cukurova University Hospital: Ahmet Rencuzogullari, Melik Kagan Aktas,	Murat Aba, Ahmet Onur Demirel, Ismail Cem Eray, Burak Aydogan
22. University of Health Sciences, Adana City Training and Research Hospital: Suleyman Cetinkunar, Kemal Yener, Alper Sozutek, Oktay Irkorucu
23. Adana Ortadogu Private Hospital: Mehmet Bayrak, Yasemin Altintas, Omer Alabaz
24. Kahramanmaras Afsin District State Hospital: Ahmet Atasever, Guven Erdogrul
25. Necip Fazil City Hospital: Aydın Hakan Kupeli, Bahtiyar Muhammedoglu, Suleyman Kokdas, Murat Kaya, Erkan Uysal
26. University of Health Sciences, Kutahya Evliya Celebi Training and Research Hospital: Ali Cihat Yildirim, Sezgin Zeren, Mehmet Fatih Ekici, Mustafa Cem Algin
27. University of Health Sciences, Samsun Training and Research Hospital: Gultekin Ozan Kucuk, Huseyin Eraslan, Engin Aybar, Suleyman Polat, Alper Ceylan
28. Uludag University Hospital: Ozgen Isik, Said Kural, Ahmet Aktas, Burak Bakar
29. Kestel State Hospital: Mustafa Yener Uzunoglu
30. Medicana Bursa Hospital: Baris Gulcu, Ersin Ozturk, Ali Onder Devay, Ersoy Taspinar
31. Bursa City Hospital-1: Ozkan Balcin, Fuat Aksoy, Gokhan Garip
32. Bursa City Hospital-2: Omer Yalkin, Nidal Iflazoglu, Direnc Yigit, Rumeysa Betul Kaya
33. Hatay Mustafa Kemal University Hospital: Mustafa Ugur, Erol Kilic, Akin Dedemoglu, Rasim Ersin Arslan, Muhyittin Temiz
34. University of Health Sciences, Tepecik Training and Research Hospital: Cengiz Aydin, Semra Demirli Atici, Tayfun Kaya, Selen Ozturk, Bulent Calik, Gizem Kilinc
35. Izmir Katip Celebi University Ataturk Training and Research Hospital: Erdinc Kamer, Turan Acar, Nihan Acar, Fevzi Cengiz
36. University of Health Sciences, Izmir Bozyaka Training and Research Hospital: Orhan Ureyen, Sedat Tan, Mehmet Yildirim, Enver Ilhan
37. Bakırçay University Cigli Training and Research Hospital: Yigit Turk
38. Urla State Hospital: Ahmet Turan Durak
39. Buca Seyfi Demirsoy Training and Research Hospital: Mehmet Yilmaz, Metin Mercan, Recep Atci
40. Dokuz Eylul University Hospital: Selman Sokmen, Tayfun Bisgin, Tufan Egeli, Yasemin Yildirim, Turugsan Safak
41. Tire State Hospital: Kazim Celik
42. Aydın Adnan Menderes University Hospital: Eyup Murat Yilmaz, Mahir Kirnap, Ahmet Ender Demirkiran, Ulas Utku Sekerci
43. Aydin State Hospital: Erkan Karacan
44. Didim State Hospital: Ethem Bilgic
45. Liv Hospital Ankara: Mehmet Mahir Ozmen, Cem Emir Guldogan, Emre Gundogdu, Munevver Moran
46. Hacettepe University Hospital: Timucin Erol, Hilmi Anil Dincer, Busenur Kirimtay, Sumeyye Yilmaz, Omer Cennet
47. Yildirim Beyazit University Yenimahalle Training and Research Hospital: Alp Yildiz, Aybala Yildiz, Can Sahin
48. Ankara University Ibni Sina Hospital: Cihangir Akyol, Mehmet Ali Koc, Siyar Ersoz, Anil Turhan, Can Konca
49. Baskent University Ankara Hospital: Tugan Tezcaner, Murathan Erkent, H. Onur Aydin, Tevfik Avci
50. Ankara Training and Research Hospital: Saygin Altiner
51. Memorial Ankara Hospital: Igbal Osmanov
52. Sincan State Hospital: Ahmet Cihangir Emral, Gokay Cetinkaya
53. Gulhane Training and Research Hospital: Emin Lapsekili, Merve Sakca, Sebnem Cimen, Dogan Ozen, Erdem Baran Kozan
54. Ankara Oncology Training and Research Hospital: Lutfi Dogan, Elifcan Haberal, Bengi Balci, Okan Kayhan, Bulent Aksel
55. University of Health Sciences, Ankara Diskapi Yildirim Beyazid Training and Research Hospital: Harun Karabacak, Cem Azili, Faruk Yazici, Muhammed Apaydin, Ismail Oskay Kaya
56. Ankara City Hospital: Erdinc Cetinkaya, Tezcan Akin, Gizem Gunes, Huseyin Turap, Deniz Aslan, Ali Eba Demirbag
57. Gazi University Hospital: Basak Bolukbasi, Berkay Enes Karaca, Ece Ozturk, Elif Ozeller, Gulsum Sueda Kayacan
58. Selcuk University Hospital: Ilhan Ece, Serdar Yormaz, Bayram Colak, Akin	Calisir, Mustafa Sahin
59. Konya City Hospital: Kemal Arslan, Ismail Hasirci, Kemal Arslan, Mehmet Esref Ulutas, Sukru Hakan Metin
60. Zonguldak Bulent Ecevit University Hospital: Fatma Ayca Gultekin
61. Elazig Fethi Sekin City Hospital: Zeynep Ozkan, Onur Ilhan, Tamer Gundogdu, Rumeysa Kevser Liman, Burhan Hakan Kanat
62. University of Health Sciences, Trabzon Kanuni Training and Research Hospital: Altan Aydin
63. Pamukkale University Hospital: Ugur Sungurtekin, Utku Ozgen, Muhammed Rasid Aykota
64. Sakarya University Training and Research Hospital: Fatih Altintoprak, Emre Gonullu, Guner Cakmak, Ugur Can Dulger, Baris Mantoglu, Hakan Demir, Emrah Akin
65. Toyotasa Emergency Hospital: Erhan Eroz
66. Mugla Sitki Kocman University Training and Research Hospital: Okay Nazli, Ozcan Dere, Mustafa Aykut Dadasoglu
67. Manisa Celal Bayar University Hospital: Eray Kara, Semra Tutcu, Ilhami Solak, Ilayda Gencer, Alperen Dalkiran
68. Usak Training and Research Hospital: Baris Sevinc, Omer Karahan, Nurullah Damburaci
69. Bandirma State Hospital: Erdem Sari, Tamer Akay, Alpaslan Fedayi Calta, Abdullah Ozdemir
70. Balikesir State Hospital: Nurian Ohri
71. Kirikkale Yuksek Ihtisas Hospital: Ilker Ermis
72. Ege University Hospital: Osman Bozbiyik, Murat Ozdemir, Berk Goktepe, Batuhan Demir, Ozgür Kilincarslan
73. Antalya Training and Research Hospital: Umut Riza Gunduz, Mehmet Olcum, Onur Ilkay Dincer, Remzi Can Cakir, Bulent Dinc
74. Kocaeli State Hospital: Enes Sahin, Emrah Uludag, Yusuf Arslan, Gokhan	Posteki, Ahmet Oktay
75. Kocaeli University Hospital: Ozan Can Tatar, Sertac Ata Guler, Nihat Zafer Utkan
76. Erzurum Regional Training and Research Hospital: Serkan Tayar, Yasar Copelci, Murat Kartal, Tolga Kalayci, Mustafa Yeni
77. Siirt State Hospital: Ahmet Cagri Buyukkasap
78. Giresun University Faculty of Medicine Training and Research Hospital: Selahattin Vural, Tugrul Kesicioglu, Ismail Aydin, Mehmet Gulmez, Can Saracoglu
79. Sivas Cumhuriyet University Hospital: Omer Topcu, Atilla Kurt, Sinan Soylu, Begum Kurt, Musa Serin
80. Diyarbakir Gazi Yasargil Training and Research Hospital: Salim Ilksen Basceken, Ebubekir Gundes, Mervan Savda, Ali Zeynel Abidin Balkan, Mehmet Nuri Yildiz
81. Harran University Training and Research Hospital: Ali Uzunkoy, Emre Karaca, Ahmet Berkan
82. Erzincan University Hospital: Arda Isik
83. Kastamonu Training and Research Hospital: Yasin Alper Yildiz, Zafer Ergul
84. Eskisehir Osmangazi University Hospital: Necdet Fatih Yasar, Bartu Badak, Ata Ozen, Melih Velipasaoglu, Iyimser Ure

**Acknowledgements:**

We are grateful to Tibbi Akademik for their academic contributions to this article.

We are grateful to Professor Alp Ozgun Borcek, MD at the Gazi University Faculty of Medicine, Department of Neurosurgery for the use of their servers for secure online data collection.
